# Factors associated with low school readiness, a linked health and education data study in Wales, UK

**DOI:** 10.1101/2022.08.14.22278759

**Authors:** Amrita Bandyopadhyay, Emily Marchant, Hope Jones, Michael Parker, Julie Evans, Sinead Brophy

**Affiliations:** National Centre for Population Health and Wellbeing Research, Swansea University Medical School, Wales, SA2 8PP, UK; Public Health Wales, Keir Hardie University Health Park, CF48 1BZ, Wales. UK

**Keywords:** School readiness, Educational attainment, Foundation Phase, Child health, Routine data, Data linkage, Cohort study

## Abstract

**Background:** School readiness is a measure of a child’s cognitive, social, and emotional readiness to begin formal schooling. Children with low school readiness need additional support from schools for learning, developing required social and academic skills, and catching-up with their school-ready peers. This study aims to identify the modifiable risk factors associated with low school readiness using linked routine data for children in Wales.

**Method:** This was a longitudinal data linkage cohort study. The cohort comprises of children who completed the Foundation Phase assessment between 2012 and 2018. Individuals were identified by linking Welsh Demographic Service and Pre16 Education Attainment datasets. School readiness was assessed via the binary outcome of the Foundation Phase assessment (achieved/not achieved). This study used multivariable logistic regression model and decision tree to identify and weight the most important risk factors associated with low school readiness.

**Results:** In order of importance, logistic regression identified maternal learning difficulties (adjusted odds ratio 5.35(95% confidence interval 3.97-7.22)), childhood epilepsy (2.95(2.39-3.66)), very low birth weight (2.24(1.86-2.70), being a boy (2.11(2.04-2.19)), being on free school meals (1.85(1.78–1.93)), living in most deprived area (1.67(1.57–1.77)), maternal death (1.47(1.09-1.98)), and maternal diabetes (1.46(1.23 - 1.78)) as factors associated with low school readiness. Using a decision tree, eligibility for free school meals, being a boy, absence/low attendance at school, being a younger child (e.g., August born), being born low birth weight, and not being breastfed were factors found to be associated with low school readiness

**Conclusion:** This work suggests that public health interventions focusing on children who are: boys, living in deprived areas, have poor early years attendance, have parents with learning difficulties, have parents with an illness or have illnesses themselves, would make the most difference to school readiness in the population.

## Introduction

### Background

Early childhood education shapes the direction of a child’s development enhances their ability to learn in the school environment and strengthens their foundation for lifelong learning (1,2). School readiness encompasses cognitive, social, and emotional aspects and indicates if a child can achieve at an appropriate level in formal school. School readiness is also a determinant of a child’s health and wellbeing over their life course (3,4). It is strongly linked to the child’s pre-school environment, and it indicates that the child has acquired the necessary social skills, emotional skills, knowledge, and attitude to effectively engage and learn in school. School readiness is defined by a child’s physical well-being and motor development (e.g., co-ordination, fine motor-skills), social and emotional development (co-operation, empathy, and the ability to express their emotion), approaches towards learning (enthusiasm, curiosity, temperament), language and communication (listening and speaking), basic knowledge (essential vocabulary and numbers) and cognitive skills (problem solving) (4).

A review of published literature on the risk factors associated with school readiness indicates that area-level characteristics, parental demography, and parental and child health conditions play a significant role in the school readiness of a child. Factors associated with higher school readiness include higher levels of child care provision in the area where the child is brought up (5,6), living in private housing (7), mother’s age (between late twenties or thirties) (7,8), breastfeeding (higher rates and longer duration) (7,9), child living with both parents, a nurturing parenting style (7,10) and parents with good physical (10,11) and mental health (7,9,10). Similarly, good physical health of the child (being born at term and a healthy birth weight) (12,13) is also associated with higher school readiness. Conversely, low access to childcare, higher levels of unemployment (area and family level), living in social housing and exposure to poor environment such as damp are associated with lower school readiness. Parental factors including maternal heavy drinking behaviours (14), mother who smoked during pregnancy (5,12), teenage mothers or older mothers (35+ years) and parents with poor physical health (hypertension, diabetes), poor mental health, single parent or step-parent families, low expectations by the parent for the child, preterm or low birth weight child, and poor health of the child are also associated with low school readiness (6,7).

Since being school ready is associated with the positive outcomes such as good health and wellbeing, and economic and employment prospects in later life, improving school readiness is a necessary strategy for economic development and social mobility (15). If children are not school ready, it can take many years for them to catch up with their peers, if ever, and therefore contribute to widening inequalities. School readiness has been identified as a key public health concern in a recent review of UK public health systems and policy approaches to early child development (16). Therefore, identifying the modifiable risk factors is a priority in closing the gap in children’s school readiness and improving outcomes for children throughout their life course.

### Objective

The aim of the study is to identify and weight the most significant modifiable risk factors of low school readiness using linked routine data for children in Wales. The factors which are associated with school readiness are examined using; a) traditional statistical methods (multivariable logistic regression model) and b) data driven supervised machine learning classification algorithm (decision tree).

## Method

### Sample selection and data linkage

In this cohort study, the study population was derived by linking the Welsh Demographic Service (WDS) dataset (administrative dataset about individuals in Wales that use NHS services) and the Pre16 Education Attainment dataset (individual-level administrative data relating to the education system in Wales). The study population consists of children who completed Foundation Phase (a curriculum for children aged 3-7 years) (17)between 2012 and 2018. The data linkage was done using an encrypted key known as Anonymised Linking Fields (ALF) in the Secure Anonymised Information Linkage (SAIL) Databank (18,19). Residential Anonymised Linking Fields (RALFs) is an encrypted residential address available in WDS dataset, which is also linked with a smaller geographic unit known as lower super output area (LSOA). Using ALFs, RALFs and LSOA the study population was anonymously linked with the individuals living with the child in the same household during the child’s Foundation Phase (20). Children without valid and continuous RALF in WDS and primary care records in Welsh Longitudinal General Practice (WLGP) dataset in SAIL until their completion of Foundation Phase were not included in the study to ensure the complete coverage of exposure and outcome data during the study period. The study population was linked with the National Community Child Health Database (NCCHD) to obtain birth and maternal records during childbirth. Records with missing maternal identifiers and mothers with no primary care record in WLGP dataset were not included in the study. The flow diagram of the selection of the study population is presented in Figure 1.

**Figure 1:**
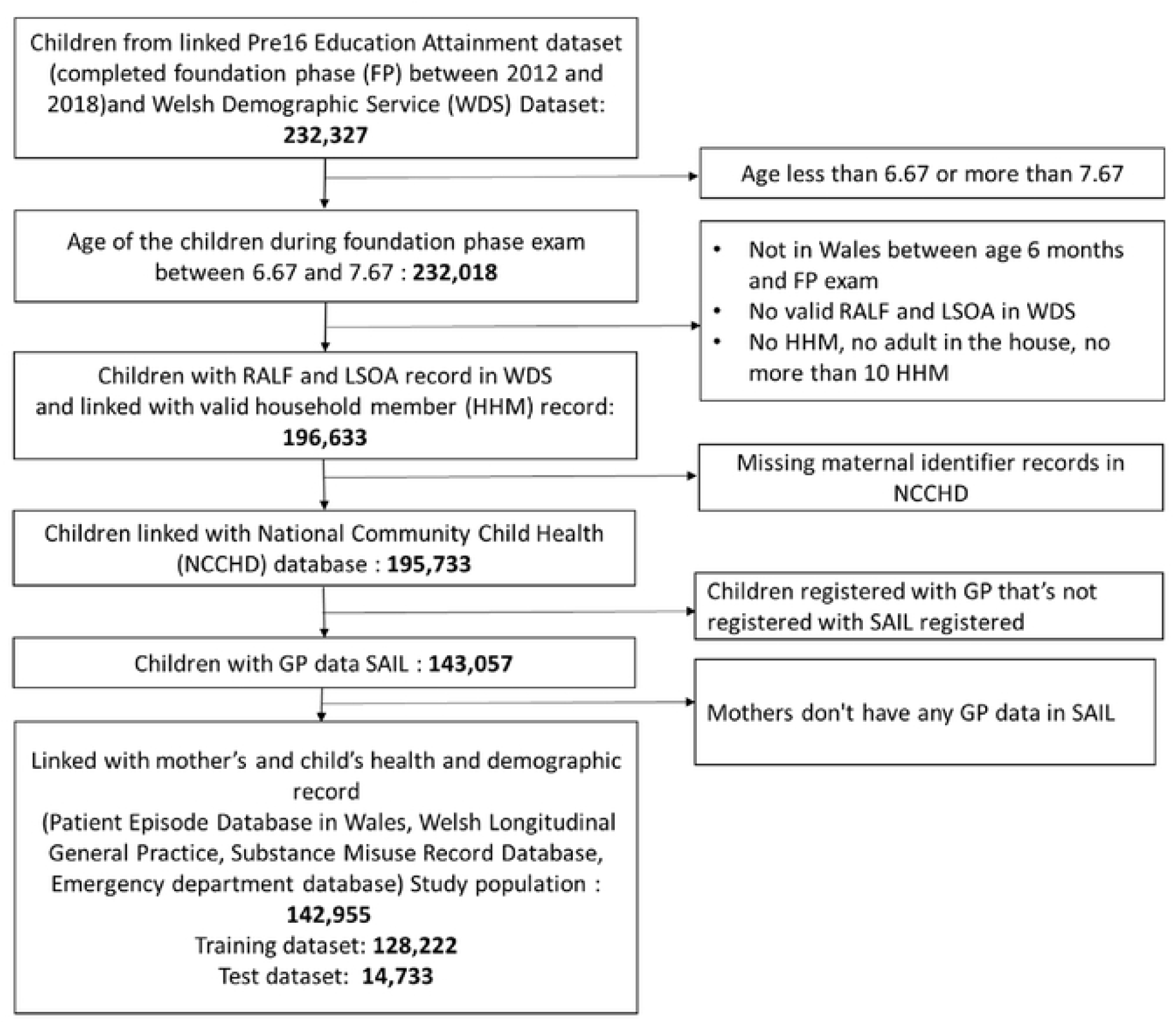
Flow diagram of the study population.

### Risk factors from routine data

In this study the selection of risk factors associated with low school readiness has been informed by the literature review undertaken at the inception of the study. The risk factors had been selected from the routinely collected electronic administrative and health datasets. General demography and birth-related variables including gender, gestational age, birth weight, breastfeeding, mode of delivery (caesarean section/assisted delivery/natural delivery) and maternal age at childbirth were obtained from NCCHD. The multiple birth (singleton/non-singleton) flag was derived using week of birth of the child, encrypted maternal identifier and the birth order of the children. To identify the children who lost their mother before the Foundation Phase, a binary variable was derived. Maternal physical health (diabetes, cancer, anaemia, hypertension, learning difficulty) and mental health (depression, anxiety, serious mental illness, medication related to anxiety/depression) related primary and secondary care records during and after pregnancy until Foundation Phase were obtained from WLGP and hospital admission dataset - Patient Episode database in Wales (PEDW). The Substance Misuse Database (SMD) was used to populate information on the mothers’ alcohol or other substance abuse related record during the study period. Mothers’ alcohol or other drug related hospital admissions and GP visits and smoking record were also obtained from PEDW and WLGP datasets. The record of physical assault related hospital admissions of mothers during or post pregnancy was obtained from PEDW. READ code version 2 and ICD10 codes have been used to identify the health records from WLGP and PEDW dataset (see the appendix). Household characteristics such as living in a single adult household, total number of adults, and total number of other children in the household were derived from the WDS dataset. In this study the eligibility for free school meals (FSM) during Foundation Phase was used to measure the family-level deprivation of the study population. The area-level deprivation was measured by the Welsh Index of Multiple Deprivation (WIMD) 2014 which provides a measure of the relative deprivation in Wales linked to LSOA (21). The local authorities and the type of local area (urban/rural) where the children were brought up during Foundation Phase were included in the study. The hospital admission and GP records of the children for epilepsy, asthma, diabetes, ear, and eye were considered as a measure of child health conditions. Any emergency hospital admission and any accident and emergency (A&E) attendance of the study population between birth and Foundation Phase were obtained from PEDW and Emergency Department dataset (EDDS). The children’s age at the completion of their Foundation Phase and the total number of days they were absent in the school during that period were obtained from Pre16 Education Attainment dataset.

### School readiness from routine data

The binary Foundation Phase Indicator variable was obtained from the Pre16 Education Attainment dataset and was used as a measure of school readiness from the routine data in the current study. The National Curriculum evaluates school readiness using Foundation Phase Indicator at the end of early year foundation stage. The Foundation Phase Indicator represents whether the child has achieved at least the expected level 5 or above in the early stage learning goals in the following areas; i) personal and social development, well-being and cultural diversity, ii) language, literacy, and communication skills – English/Welsh and iii) mathematical development (22). In this study a binary variable has been derived based on the Foundation Phase Indicator record as a measure of school readiness from routine data.

- Low school readiness = Not achieved Foundation Phase
- School readiness = Achieved Foundation Phase

### Statistical analysis

A multivariable logistic regression model was first developed to identify and weight the most important risk factors associated with school readiness. Next, we built a prediction model using supervised machined learning classifier - decision tree. Since the children with learning difficulties tend to have much higher risk of low school readiness, we filtered out the children with learning difficulties from the regression and the prediction model. The models focused on the children without learning difficulties. Data preparation including data linkage was performed on DB2 SQL platform and the statistical analysis was done in R version 4.0.3.

### Logistic Regression

To identify the most important risk factors associated with low school readiness for children without a known learning difficulty (e.g., down’s syndrome) we used multivariable logistic regression. Variables included gender, gestational age, birthweight, breastfeeding, caesarean section, multiple birth, maternal age, maternal death before Foundation Phase, maternal physical and mental health, child physical (epilepsy, asthma, diabetes, ear, and eye) and mental health conditions (depression, anxiety), FSM, local area status and number of adults and children living in the same household. The significant risk factors of low school readiness are presented with their adjusted Odds Ratio (aOR) and 95% confidence interval (CI).

### Decision Tree

Since the outcome variable was a binary variable, classification tree – decision tree algorithms were developed using RPART (Recursive Partitioning And Regression Trees) packages in R (23,24). The algorithm repeatedly partitions the data into multiple sub-spaces to reach the homogeneous end sub-space hence it is called recursive partitioning. For decision trees, the data for one representative local authority was take out from the dataset and was used as the testing dataset to validate the model performance and examine generalisability within areas of Wales.

## Results

### Overall sample characteristics

The study population consisted of 142,955 children (Training dataset: 128,222, Testing dataset: 14,733) who completed Foundation Phase between 2012 and 2018 in Wales (see Table 1). Overall, 14.32% (Training dataset: 14.15%, Testing dataset: 15.75%) of children did not achieve in Foundation Phase, indicating low school readiness. The study population consisted of 51.24% boys, 42.87% of children who were not breastfed and 24.83% of children were born via caesarean section. 8.33% of children were born to mothers aged below 19, 0.14% of mothers had learning difficulties and 0.23% of children lost their mother before their Foundation Phase assessment. There were 0.1% mothers who had alcohol related hospital admission, 0.36% with substance abuse and 14.63**%** had smoking record in WLGP during pregnancy. There were 0.64% of children with epilepsy related GP visit, 0.46% had a hospital admission record for epilepsy. For asthma 3.30% and for ear problems 4.54% of children were admitted to hospital before they completed Foundation Phase. 56.37% children had at least one emergency hospital admission and 66.4% had A&E records anytime between birth and Foundation Phase. Overall, 0.90% of children (Training dataset: 0.88%, Testing dataset 1.07%) were diagnosed with a learning difficulty. 22.05% of children were in single adult households, 19.57% of children were eligible for FSM and 25.66% lived in most deprived area measured by WIMD. Overall characteristics of the study population have been described in Table 1.

**Table 1:**
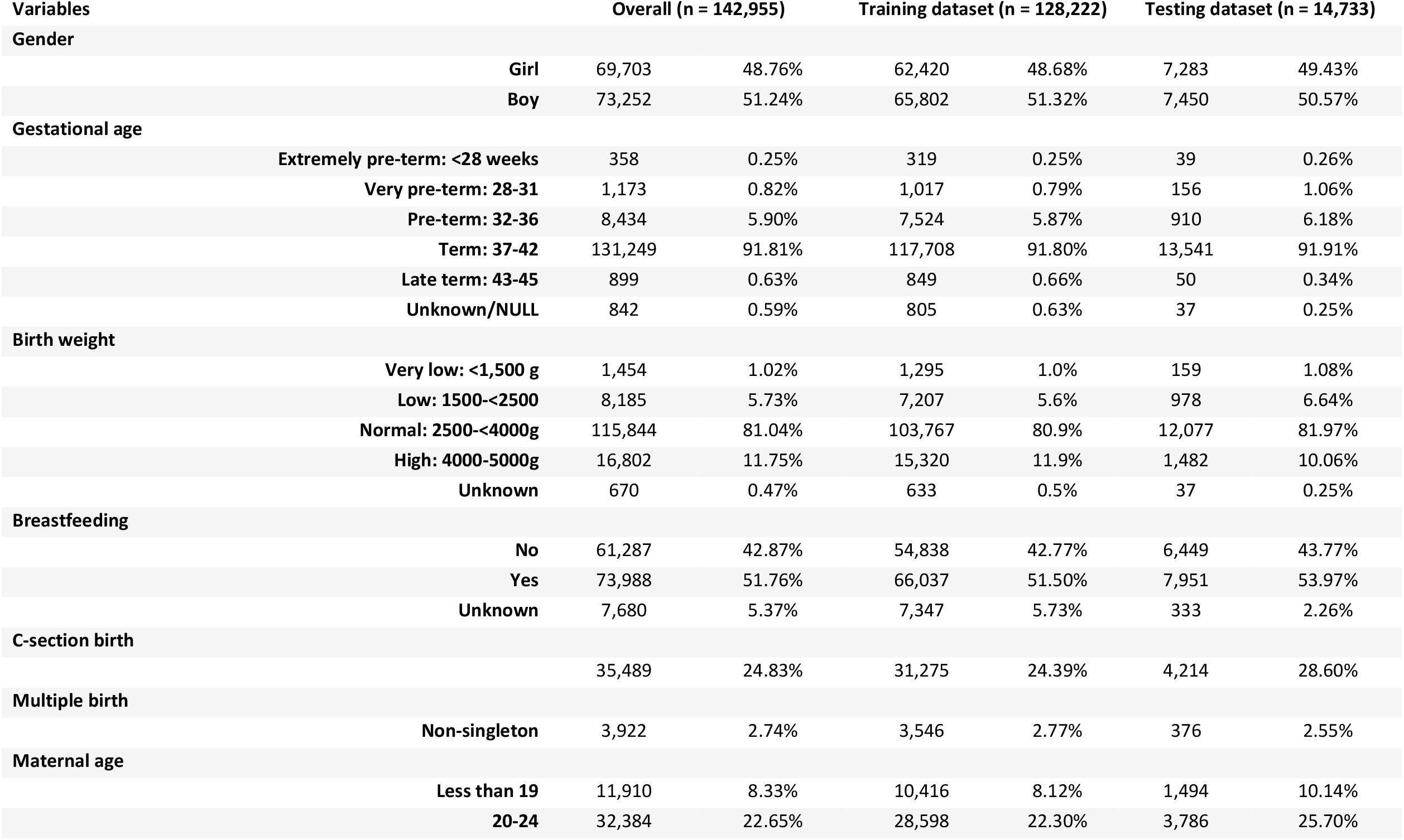
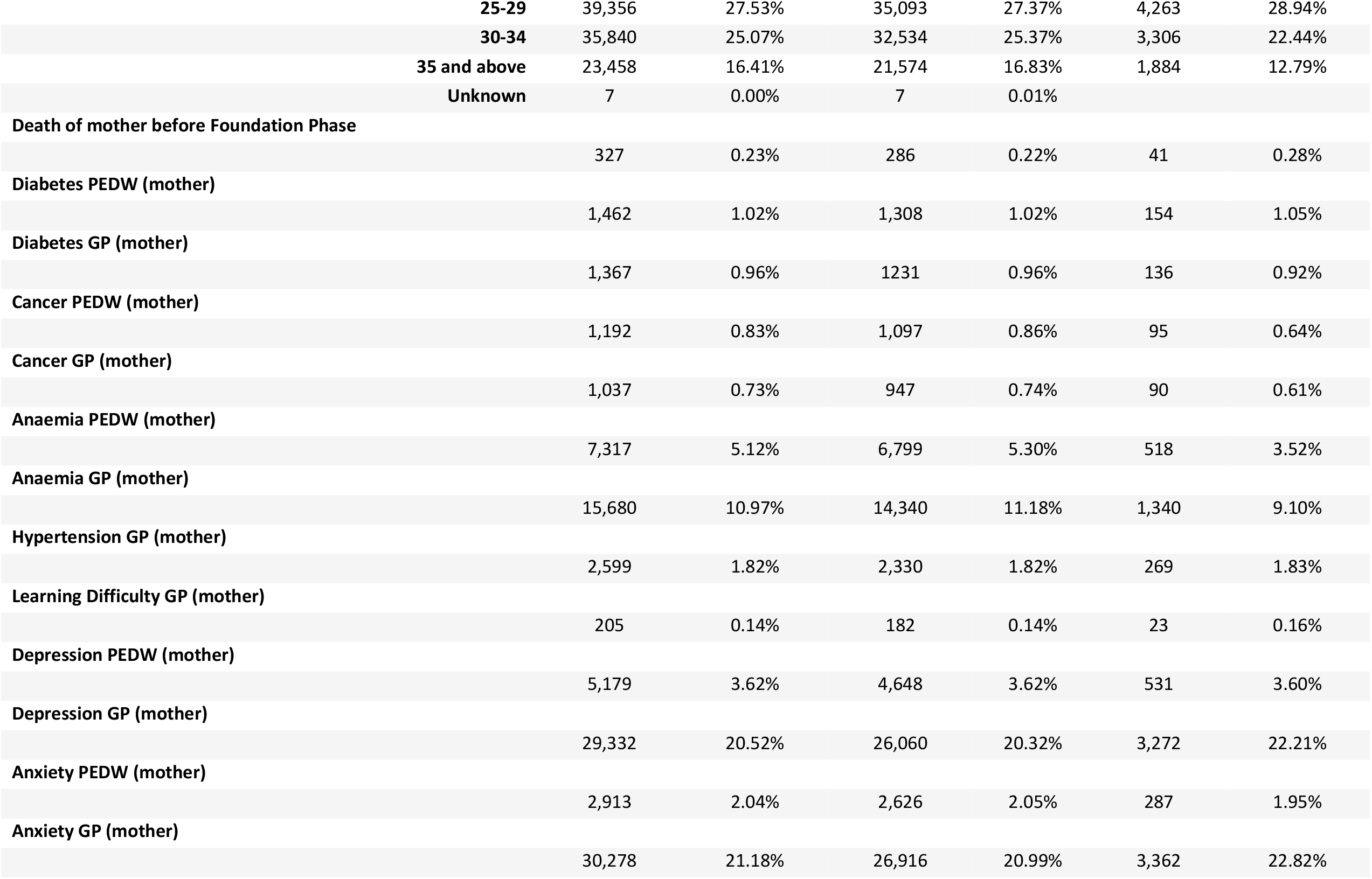
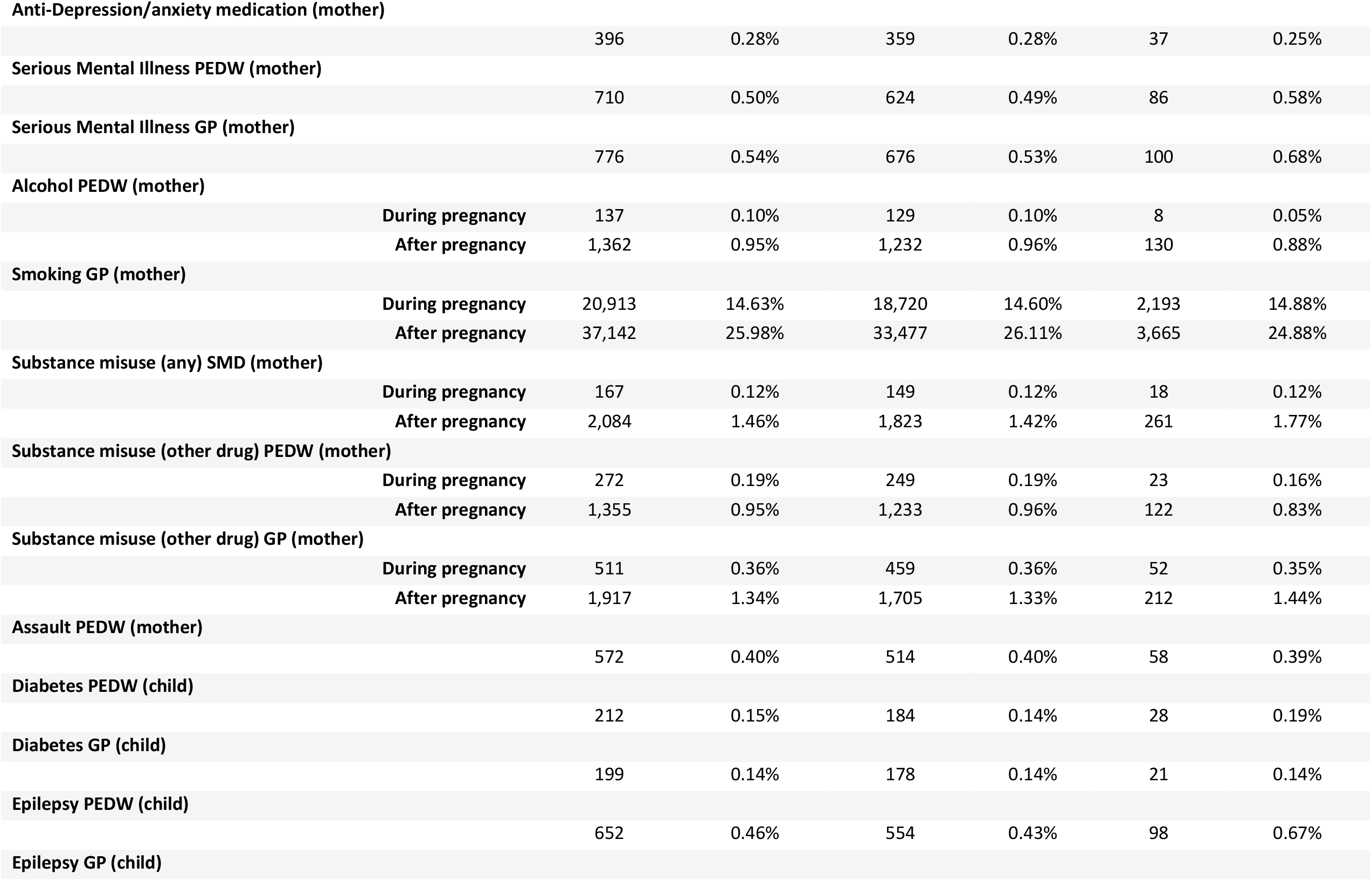
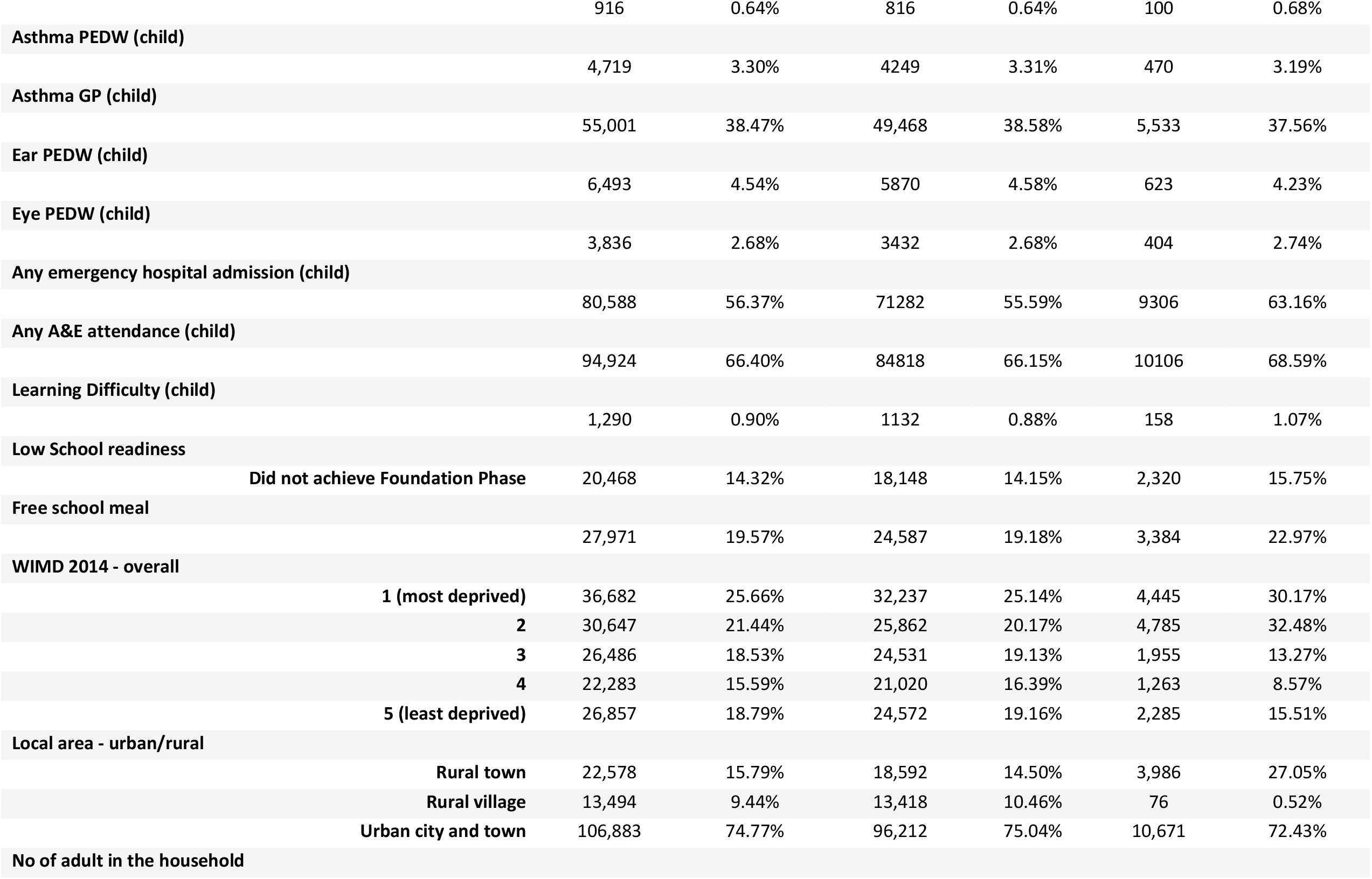
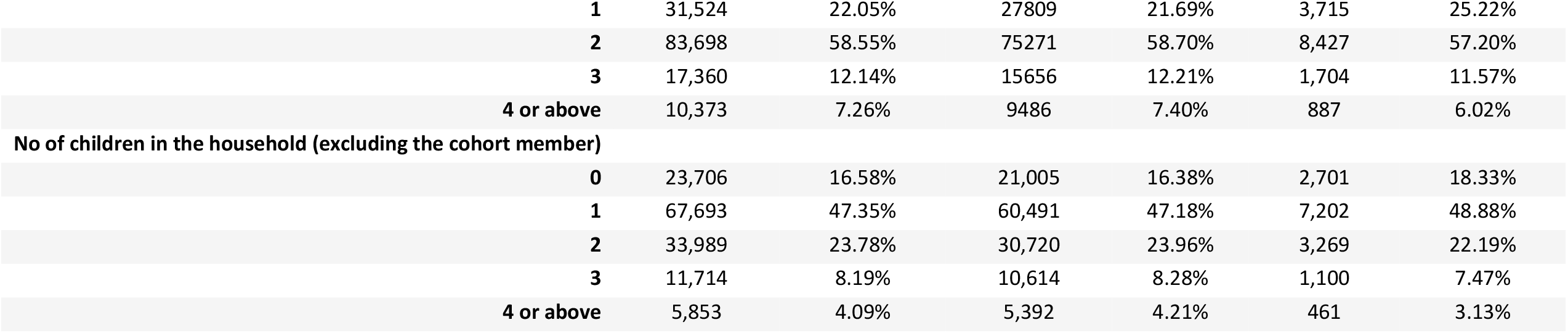
Characteristics of the study population.

| Variables |  | Overall (n = 142,955) |  | Training dataset (n = 128,222) |  | Testing dataset (n = 14,733) |  |
| --- | --- | --- | --- | --- | --- | --- | --- |
| **Gender** |  |  |  |  |  |  |  |
|  | **Girl** | 69,703 | 48.76% | 62,420 | 48.68% | 7,283 | 49.43% |
|  | **Boy** | 73,252 | 51.24% | 65,802 | 51.32% | 7,450 | 50.57% |
| **Gestational age** |  |  |  |  |  |  |  |
|  | **Extremely pre-term: <28 weeks** | 358 | 0.25% | 319 | 0.25% | 39 | 0.26% |
|  | **Very pre-term: 28-31** | 1,173 | 0.82% | 1,017 | 0.79% | 156 | 1.06% |
|  | **Pre-term: 32-36** | 8,434 | 5.90% | 7,524 | 5.87% | 910 | 6.18% |
|  | **Term: 37-42** | 131,249 | 91.81% | 117,708 | 91.80% | 13,541 | 91.91% |
|  | **Late term: 43-45** | 899 | 0.63% | 849 | 0.66% | 50 | 0.34% |
|  | **Unknown/NULL** | 842 | 0.59% | 805 | 0.63% | 37 | 0.25% |
| **Birth weight** |  |  |  |  |  |  |  |
|  | **Very low: <1,500 g** | 1,454 | 1.02% | 1,295 | 1.0% | 159 | 1.08% |
|  | **Low: 1500-<2500** | 8,185 | 5.73% | 7,207 | 5.6% | 978 | 6.64% |
|  | **Normal: 2500-<4000g** | 115,844 | 81.04% | 103,767 | 80.9% | 12,077 | 81.97% |
|  | **High: 4000-5000g** | 16,802 | 11.75% | 15,320 | 11.9% | 1,482 | 10.06% |
|  | **Unknown** | 670 | 0.47% | 633 | 0.5% | 37 | 0.25% |
| **Breastfeeding** |  |  |  |  |  |  |  |
|  | **No** | 61,287 | 42.87% | 54,838 | 42.77% | 6,449 | 43.77% |
|  | **Yes** | 73,988 | 51.76% | 66,037 | 51.50% | 7,951 | 53.97% |
|  | **Unknown** | 7,680 | 5.37% | 7,347 | 5.73% | 333 | 2.26% |
| **C-section birth** |  |  |  |  |  |  |  |
|  |  | 35,489 | 24.83% | 31,275 | 24.39% | 4,214 | 28.60% |
| **Multiple birth** |  |  |  |  |  |  |  |
|  | **Non-singleton** | 3,922 | 2.74% | 3,546 | 2.77% | 376 | 2.55% |
| **Maternal age** |  |  |  |  |  |  |  |
|  | **Less than 19** | 11,910 | 8.33% | 10,416 | 8.12% | 1,494 | 10.14% |
|  | **20-24** | 32,384 | 22.65% | 28,598 | 22.30% | 3,786 | 25.70% |
|  | **25-29** | 39,356 | 27.53% | 35,093 | 27.37% | 4,263 | 28.94% |
|  | **30-34** | 35,840 | 25.07% | 32,534 | 25.37% | 3,306 | 22.44% |
|  | **35 and above** | 23,458 | 16.41% | 21,574 | 16.83% | 1,884 | 12.79% |
|  | **Unknown** | 7 | 0.00% | 7 | 0.01% |  |  |
| **Death of mother before Foundation Phase** |  |  |  |  |  |  |  |
|  |  | 327 | 0.23% | 286 | 0.22% | 41 | 0.28% |
| **Diabetes PEDW (mother)** |  |  |  |  |  |  |  |
|  |  | 1,462 | 1.02% | 1,308 | 1.02% | 154 | 1.05% |
| **Diabetes GP (mother)** |  |  |  |  |  |  |  |
|  |  | 1,367 | 0.96% | 1231 | 0.96% | 136 | 0.92% |
| **Cancer PEDW (mother)** |  |  |  |  |  |  |  |
|  |  | 1,192 | 0.83% | 1,097 | 0.86% | 95 | 0.64% |
| **Cancer GP (mother)** |  |  |  |  |  |  |  |
|  |  | 1,037 | 0.73% | 947 | 0.74% | 90 | 0.61% |
| **Anaemia PEDW (mother)** |  |  |  |  |  |  |  |
|  |  | 7,317 | 5.12% | 6,799 | 5.30% | 518 | 3.52% |
| **Anaemia GP (mother)** |  |  |  |  |  |  |  |
|  |  | 15,680 | 10.97% | 14,340 | 11.18% | 1,340 | 9.10% |
| **Hypertension GP (mother)** |  |  |  |  |  |  |  |
|  |  | 2,599 | 1.82% | 2,330 | 1.82% | 269 | 1.83% |
| **Learning Difficulty GP (mother)** |  |  |  |  |  |  |  |
|  |  | 205 | 0.14% | 182 | 0.14% | 23 | 0.16% |
| **Depression PEDW (mother)** |  |  |  |  |  |  |  |
|  |  | 5,179 | 3.62% | 4,648 | 3.62% | 531 | 3.60% |
| **Depression GP (mother)** |  |  |  |  |  |  |  |
|  |  | 29,332 | 20.52% | 26,060 | 20.32% | 3,272 | 22.21% |
| **Anxiety PEDW (mother)** |  |  |  |  |  |  |  |
|  |  | 2,913 | 2.04% | 2,626 | 2.05% | 287 | 1.95% |
| **Anxiety GP (mother)** |  |  |  |  |  |  |  |
|  |  | 30,278 | 21.18% | 26,916 | 20.99% | 3,362 | 22.82% |
| **Anti-Depression/anxiety medication (mother)** |  | 396 | 0.28% | 359 | 0.28% | 37 | 0.25% |
| **Serious Mental Illness PEDW (mother)** |  | 710 | 0.50% | 624 | 0.49% | 86 | 0.58% |
| **Serious Mental Illness GP (mother)** |  | 776 | 0.54% | 676 | 0.53% | 100 | 0.68% |
| **Alcohol PEDW (mother)** |  |  |  |  |  |  |  |
|  | **During pregnancy** | 137 | 0.10% | 129 | 0.10% | 8 | 0.05% |
|  | **After pregnancy** | 1,362 | 0.95% | 1,232 | 0.96% | 130 | 0.88% |
| **Smoking GP (mother)** |  |  |  |  |  |  |  |
|  | **During pregnancy** | 20,913 | 14.63% | 18,720 | 14.60% | 2,193 | 14.88% |
|  | **After pregnancy** | 37,142 | 25.98% | 33,477 | 26.11% | 3,665 | 24.88% |
| **Substance misuse (any) SMD (mother)** |  |  |  |  |  |  |  |
|  | **During pregnancy** | 167 | 0.12% | 149 | 0.12% | 18 | 0.12% |
|  | **After pregnancy** | 2,084 | 1.46% | 1,823 | 1.42% | 261 | 1.77% |
| **Substance misuse (other drug) PEDW (mother)** |  |  |  |  |  |  |  |
|  | **During pregnancy** | 272 | 0.19% | 249 | 0.19% | 23 | 0.16% |
|  | **After pregnancy** | 1,355 | 0.95% | 1,233 | 0.96% | 122 | 0.83% |
| **Substance misuse (other drug) GP (mother)** |  |  |  |  |  |  |  |
|  | **During pregnancy** | 511 | 0.36% | 459 | 0.36% | 52 | 0.35% |
|  | **After pregnancy** | 1,917 | 1.34% | 1,705 | 1.33% | 212 | 1.44% |
| **Assault PEDW (mother)** |  | 572 | 0.40% | 514 | 0.40% | 58 | 0.39% |
| **Diabetes PEDW (child)** |  | 212 | 0.15% | 184 | 0.14% | 28 | 0.19% |
| **Diabetes GP (child)** |  | 199 | 0.14% | 178 | 0.14% | 21 | 0.14% |
| **Epilepsy PEDW (child)** |  | 652 | 0.46% | 554 | 0.43% | 98 | 0.67% |
| **Epilepsy GP (child)** |  |  |  |  |  |  |  |
|  |  | 916 | 0.64% | 816 | 0.64% | 100 | 0.68% |
| **Asthma PEDW (child)** |  |  |  |  |  |  |  |
|  |  | 4,719 | 3.30% | 4249 | 3.31% | 470 | 3.19% |
| **Asthma GP (child)** |  |  |  |  |  |  |  |
|  |  | 55,001 | 38.47% | 49,468 | 38.58% | 5,533 | 37.56% |
| **Ear PEDW (child)** |  |  |  |  |  |  |  |
|  |  | 6,493 | 4.54% | 5870 | 4.58% | 623 | 4.23% |
| **Eye PEDW (child)** |  |  |  |  |  |  |  |
|  |  | 3,836 | 2.68% | 3432 | 2.68% | 404 | 2.74% |
| **Any emergency hospital admission (child)** |  |  |  |  |  |  |  |
|  |  | 80,588 | 56.37% | 71282 | 55.59% | 9306 | 63.16% |
| **Any A&E attendance (child)** |  |  |  |  |  |  |  |
|  |  | 94,924 | 66.40% | 84818 | 66.15% | 10106 | 68.59% |
| **Learning Difficulty (child)** |  |  |  |  |  |  |  |
|  |  | 1,290 | 0.90% | 1132 | 0.88% | 158 | 1.07% |
| **Low School readiness** |  |  |  |  |  |  |  |
|  | **Did not achieve Foundation Phase** | 20,468 | 14.32% | 18,148 | 14.15% | 2,320 | 15.75% |
| **Free school meal** |  |  |  |  |  |  |  |
|  |  | 27,971 | 19.57% | 24,587 | 19.18% | 3,384 | 22.97% |
| **WIMD 2014 - overall** |  |  |  |  |  |  |  |
|  | **1 (most deprived)** | 36,682 | 25.66% | 32,237 | 25.14% | 4,445 | 30.17% |
|  | **2** | 30,647 | 21.44% | 25,862 | 20.17% | 4,785 | 32.48% |
|  | **3** | 26,486 | 18.53% | 24,531 | 19.13% | 1,955 | 13.27% |
|  | **4** | 22,283 | 15.59% | 21,020 | 16.39% | 1,263 | 8.57% |
|  | **5 (least deprived)** | 26,857 | 18.79% | 24,572 | 19.16% | 2,285 | 15.51% |
| **Local area - urban/rural** |  |  |  |  |  |  |  |
|  | **Rural town** | 22,578 | 15.79% | 18,592 | 14.50% | 3,986 | 27.05% |
|  | **Rural village** | 13,494 | 9.44% | 13,418 | 10.46% | 76 | 0.52% |
|  | **Urban city and town** | 106,883 | 74.77% | 96,212 | 75.04% | 10,671 | 72.43% |
| **No of adult in the household** |  |  |  |  |  |  |  |

|  |  |  |  |  |  |  |
| --- | --- | --- | --- | --- | --- | --- |
| **1** | 31,524 | 22.05% | 27809 | 21.69% | 3,715 | 25.22% |
| **2** | 83,698 | 58.55% | 75271 | 58.70% | 8,427 | 57.20% |
| **3** | 17,360 | 12.14% | 15656 | 12.21% | 1,704 | 11.57% |
| **4 or above** | 10,373 | 7.26% | 9486 | 7.40% | 887 | 6.02% |
| **No of children in the household (excluding the cohort member)** |  |  |  |  |  |  |
| **0** | 23,706 | 16.58% | 21,005 | 16.38% | 2,701 | 18.33% |
| **1** | 67,693 | 47.35% | 60,491 | 47.18% | 7,202 | 48.88% |
| **2** | 33,989 | 23.78% | 30,720 | 23.96% | 3,269 | 22.19% |
| **3** | 11,714 | 8.19% | 10,614 | 8.28% | 1,100 | 7.47% |
| **4 or above** | 5,853 | 4.09% | 5,392 | 4.21% | 461 | 3.13% |

### Logistic regression results

Significant risk factors associated with low school readiness included: maternal learning difficulty (aOR (95% CI): 5.35 (3.97 - 7.22)), child epilepsy (2.95 (2.39 - 3.66)), having a very low birthweight (2.24 (1.86 - 2.70)), boys (2.11 (2.04 - 2.19)), being eligible for FSM (1.85 (1.78 – 1.93)), being extremely preterm (1.41 (1.04 - 1.91)), living in most deprived area (1.67 (1.57 – 1.77)), and other factors, such as not being breastfed (1.25 (1.21 -1.30)), maternal death (1.47 (1.09 - 1.98)), maternal diabetes (1.46 (1.23 - 1.78)), smoking in pregnancy (1.36 (1.30- 1.43)), child hospital admissions/illness for asthma (1.12 (1.03 - 1.22)), ear (1.36 (1.26 - 1.45)) and eye (1.30 (1.18 - 1.42)), single adult household (1.08 (1.04 - 1.12)), many other children (1.63 (1.52 - 1.75)) in the household. The risk factors with their OR and upper and lower CI are presented in Table 2.

**Table 2:**
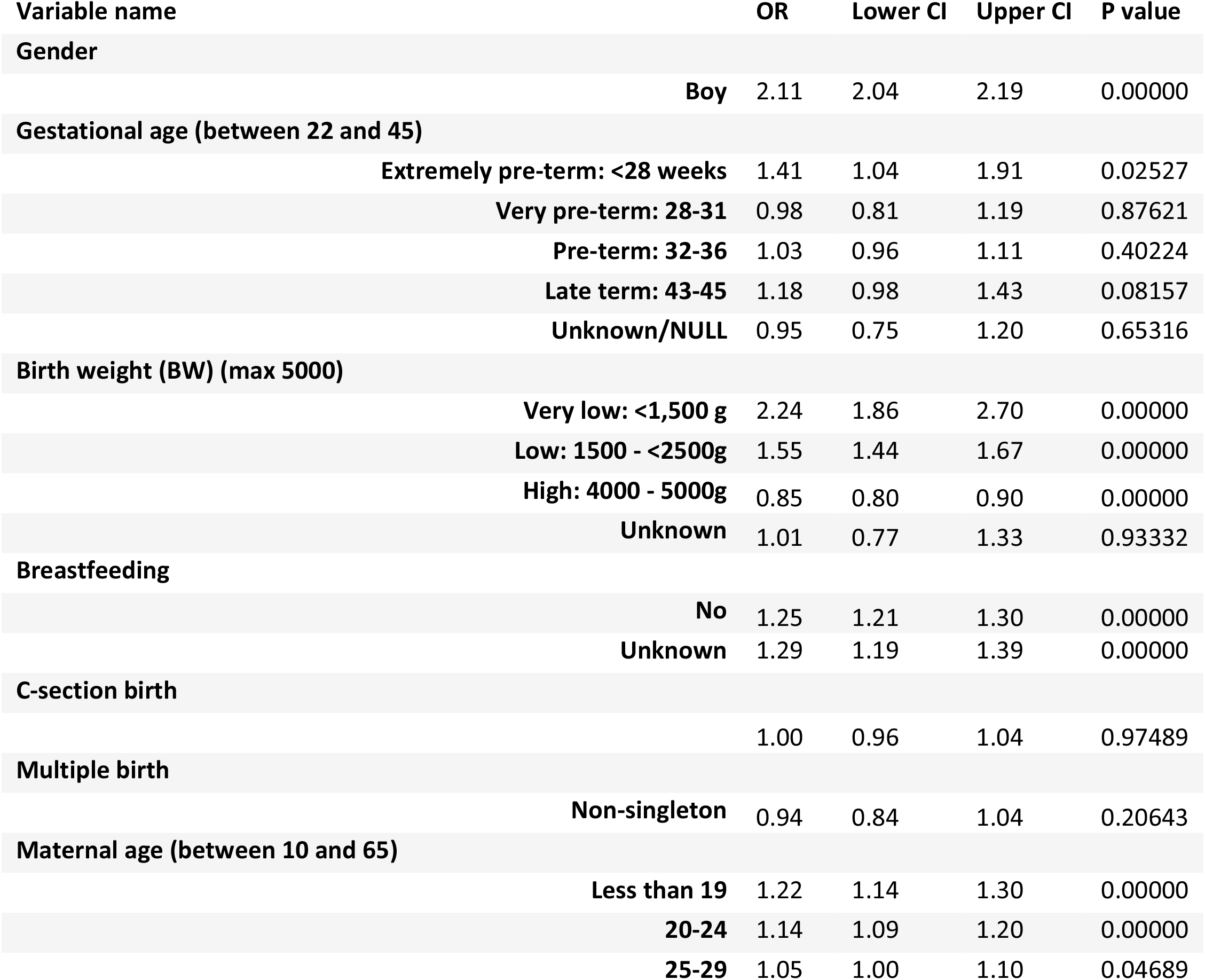
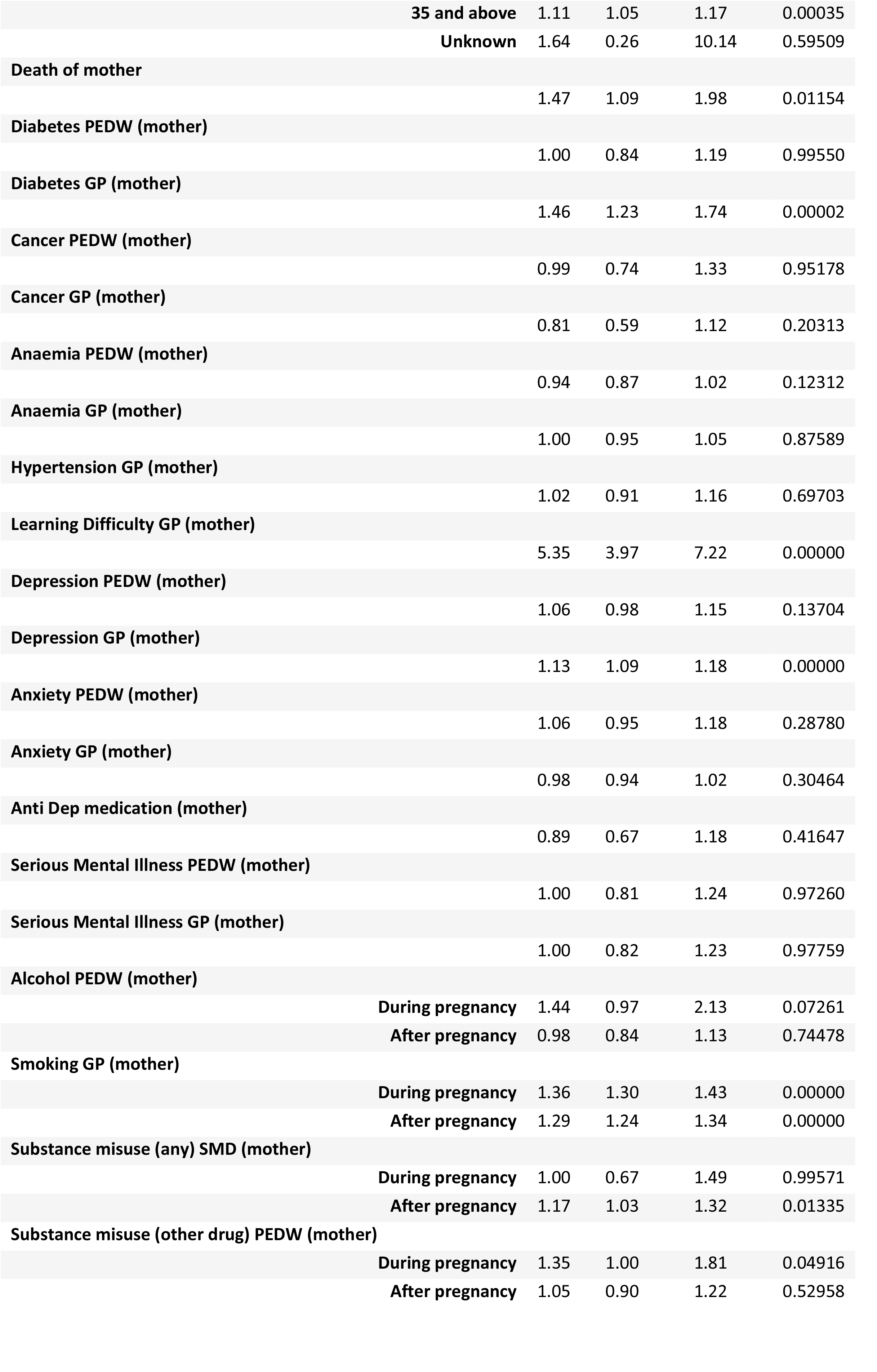
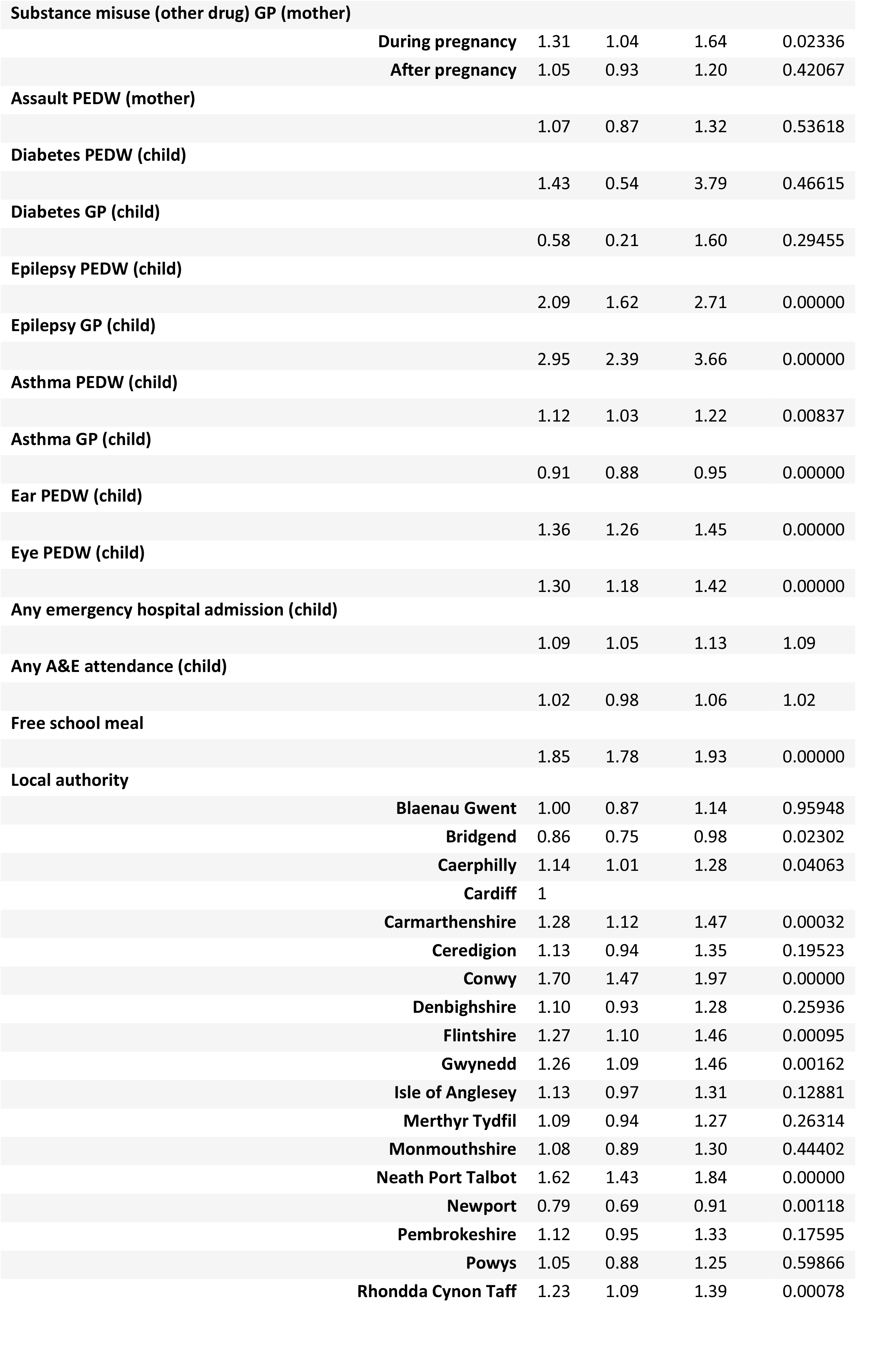
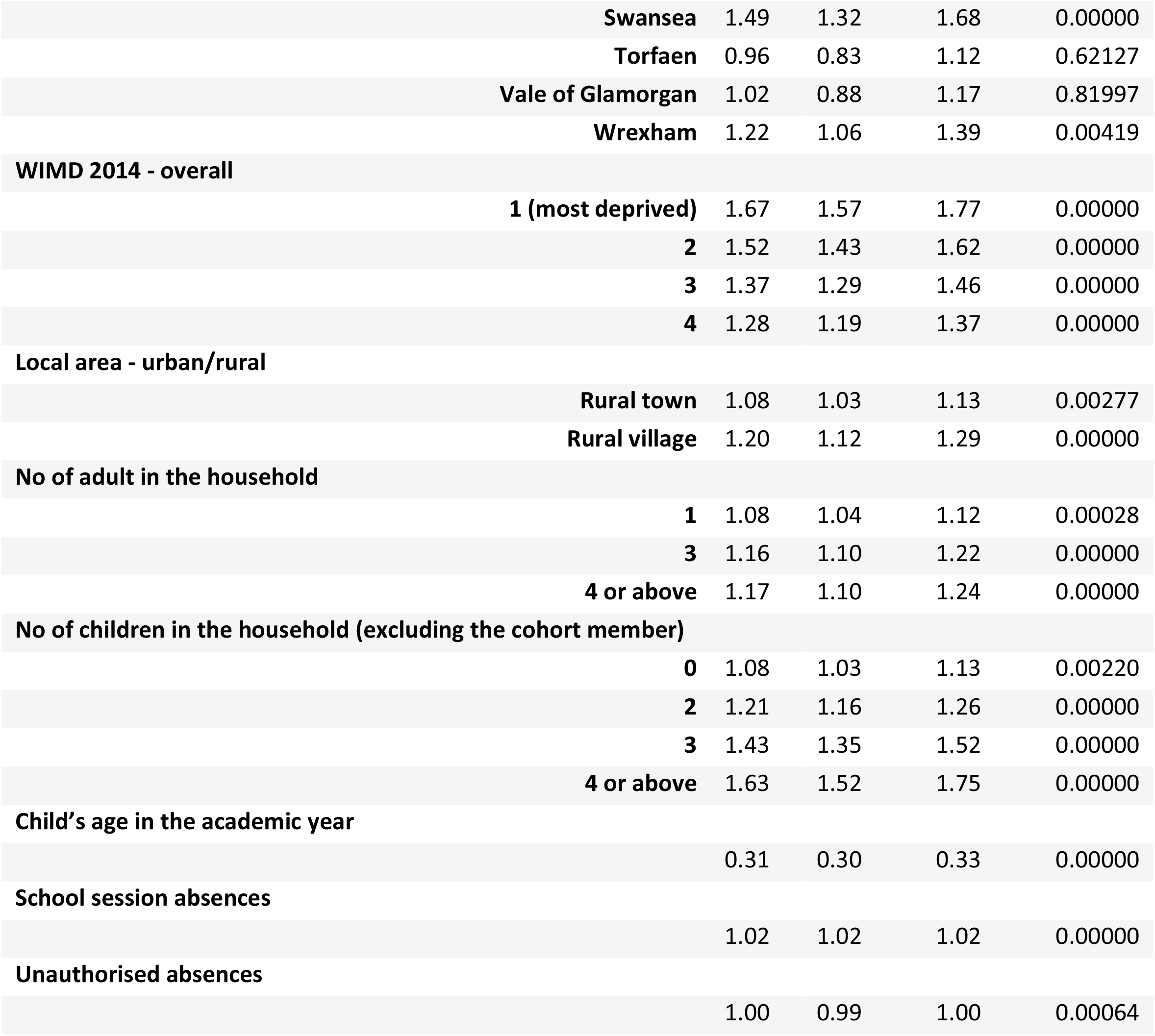
Logistic regression model to identify the risk factors associated with low school readiness.

| Variable name | OR | Lower CI | Upper CI | P value |
| --- | --- | --- | --- | --- |
| **Gender** |  |  |  |  |
| Boy | 2.11 | 2.04 | 2.19 | 0.00000 |
| **Gestational age (between 22 and 45)** |  |  |  |  |
| Extremely pre-term: <28 weeks | 1.41 | 1.04 | 1.91 | 0.02527 |
| Very pre-term: 28-31 | 0.98 | 0.81 | 1.19 | 0.87621 |
| Pre-term: 32-36 | 1.03 | 0.96 | 1.11 | 0.40224 |
| Late term: 43-45 | 1.18 | 0.98 | 1.43 | 0.08157 |
| Unknown/NULL | 0.95 | 0.75 | 1.20 | 0.65316 |
| **Birth weight (BW) (max 5000)** |  |  |  |  |
| Very low: <1,500 g | 2.24 | 1.86 | 2.70 | 0.00000 |
| Low: 1500 - <2500g | 1.55 | 1.44 | 1.67 | 0.00000 |
| High: 4000 - 5000g | 0.85 | 0.80 | 0.90 | 0.00000 |
| Unknown | 1.01 | 0.77 | 1.33 | 0.93332 |
| **Breastfeeding** |  |  |  |  |
| No | 1.25 | 1.21 | 1.30 | 0.00000 |
| Unknown | 1.29 | 1.19 | 1.39 | 0.00000 |
| **C-section birth** |  |  |  |  |
|  | 1.00 | 0.96 | 1.04 | 0.97489 |
| **Multiple birth** |  |  |  |  |
| Non-singleton | 0.94 | 0.84 | 1.04 | 0.20643 |
| **Maternal age (between 10 and 65)** |  |  |  |  |
| Less than 19 | 1.22 | 1.14 | 1.30 | 0.00000 |
| 20-24 | 1.14 | 1.09 | 1.20 | 0.00000 |
| 25-29 | 1.05 | 1.00 | 1.10 | 0.04689 |

|  |  |  |  |  |  |
| --- | --- | --- | --- | --- | --- |
|  | **35 and above** | 1.11 | 1.05 | 1.17 | 0.00035 |
|  | **Unknown** | 1.64 | 0.26 | 10.14 | 0.59509 |
| **Death of mother** |  |  |  |  |  |
|  |  | 1.47 | 1.09 | 1.98 | 0.01154 |
| **Diabetes PEDW (mother)** |  |  |  |  |  |
|  |  | 1.00 | 0.84 | 1.19 | 0.99550 |
| **Diabetes GP (mother)** |  |  |  |  |  |
|  |  | 1.46 | 1.23 | 1.74 | 0.00002 |
| **Cancer PEDW (mother)** |  |  |  |  |  |
|  |  | 0.99 | 0.74 | 1.33 | 0.95178 |
| **Cancer GP (mother)** |  |  |  |  |  |
|  |  | 0.81 | 0.59 | 1.12 | 0.20313 |
| **Anaemia PEDW (mother)** |  |  |  |  |  |
|  |  | 0.94 | 0.87 | 1.02 | 0.12312 |
| **Anaemia GP (mother)** |  |  |  |  |  |
|  |  | 1.00 | 0.95 | 1.05 | 0.87589 |
| **Hypertension GP (mother)** |  |  |  |  |  |
|  |  | 1.02 | 0.91 | 1.16 | 0.69703 |
| **Learning Difficulty GP (mother)** |  |  |  |  |  |
|  |  | 5.35 | 3.97 | 7.22 | 0.00000 |
| **Depression PEDW (mother)** |  |  |  |  |  |
|  |  | 1.06 | 0.98 | 1.15 | 0.13704 |
| **Depression GP (mother)** |  |  |  |  |  |
|  |  | 1.13 | 1.09 | 1.18 | 0.00000 |
| **Anxiety PEDW (mother)** |  |  |  |  |  |
|  |  | 1.06 | 0.95 | 1.18 | 0.28780 |
| **Anxiety GP (mother)** |  |  |  |  |  |
|  |  | 0.98 | 0.94 | 1.02 | 0.30464 |
| **Anti Dep medication (mother)** |  |  |  |  |  |
|  |  | 0.89 | 0.67 | 1.18 | 0.41647 |
| **Serious Mental Illness PEDW (mother)** |  |  |  |  |  |
|  |  | 1.00 | 0.81 | 1.24 | 0.97260 |
| **Serious Mental Illness GP (mother)** |  |  |  |  |  |
|  |  | 1.00 | 0.82 | 1.23 | 0.97759 |
| **Alcohol PEDW (mother)** |  |  |  |  |  |
|  | **During pregnancy** | 1.44 | 0.97 | 2.13 | 0.07261 |
|  | **After pregnancy** | 0.98 | 0.84 | 1.13 | 0.74478 |
| **Smoking GP (mother)** |  |  |  |  |  |
|  | **During pregnancy** | 1.36 | 1.30 | 1.43 | 0.00000 |
|  | **After pregnancy** | 1.29 | 1.24 | 1.34 | 0.00000 |
| **Substance misuse (any) SMD (mother)** |  |  |  |  |  |
|  | **During pregnancy** | 1.00 | 0.67 | 1.49 | 0.99571 |
|  | **After pregnancy** | 1.17 | 1.03 | 1.32 | 0.01335 |
| **Substance misuse (other drug) PEDW (mother)** |  |  |  |  |  |
|  | **During pregnancy** | 1.35 | 1.00 | 1.81 | 0.04916 |
|  | **After pregnancy** | 1.05 | 0.90 | 1.22 | 0.52958 |
| **Substance misuse (other drug) GP (mother)** |  |  |  |  |  |
|  | **During pregnancy** | 1.31 | 1.04 | 1.64 | 0.02336 |
|  | **After pregnancy** | 1.05 | 0.93 | 1.20 | 0.42067 |
| **Assault PEDW (mother)** |  |  |  |  |  |
|  |  | 1.07 | 0.87 | 1.32 | 0.53618 |
| **Diabetes PEDW (child)** |  |  |  |  |  |
|  |  | 1.43 | 0.54 | 3.79 | 0.46615 |
| **Diabetes GP (child)** |  |  |  |  |  |
|  |  | 0.58 | 0.21 | 1.60 | 0.29455 |
| **Epilepsy PEDW (child)** |  |  |  |  |  |
|  |  | 2.09 | 1.62 | 2.71 | 0.00000 |
| **Epilepsy GP (child)** |  |  |  |  |  |
|  |  | 2.95 | 2.39 | 3.66 | 0.00000 |
| **Asthma PEDW (child)** |  |  |  |  |  |
|  |  | 1.12 | 1.03 | 1.22 | 0.00837 |
| **Asthma GP (child)** |  |  |  |  |  |
|  |  | 0.91 | 0.88 | 0.95 | 0.00000 |
| **Ear PEDW (child)** |  |  |  |  |  |
|  |  | 1.36 | 1.26 | 1.45 | 0.00000 |
| **Eye PEDW (child)** |  |  |  |  |  |
|  |  | 1.30 | 1.18 | 1.42 | 0.00000 |
| **Any emergency hospital admission (child)** |  |  |  |  |  |
|  |  | 1.09 | 1.05 | 1.13 | 1.09 |
| **Any A&E attendance (child)** |  |  |  |  |  |
|  |  | 1.02 | 0.98 | 1.06 | 1.02 |
| **Free school meal** |  |  |  |  |  |
|  |  | 1.85 | 1.78 | 1.93 | 0.00000 |
| **Local authority** |  |  |  |  |  |
|  | **Blaenau Gwent** | 1.00 | 0.87 | 1.14 | 0.95948 |
|  | **Bridgend** | 0.86 | 0.75 | 0.98 | 0.02302 |
|  | **Caerphilly** | 1.14 | 1.01 | 1.28 | 0.04063 |
|  | **Cardiff** | 1 |  |  |  |
|  | **Carmarthenshire** | 1.28 | 1.12 | 1.47 | 0.00032 |
|  | **Ceredigion** | 1.13 | 0.94 | 1.35 | 0.19523 |
|  | **Conwy** | 1.70 | 1.47 | 1.97 | 0.00000 |
|  | **Denbighshire** | 1.10 | 0.93 | 1.28 | 0.25936 |
|  | **Flintshire** | 1.27 | 1.10 | 1.46 | 0.00095 |
|  | **Gwynedd** | 1.26 | 1.09 | 1.46 | 0.00162 |
|  | **Isle of Anglesey** | 1.13 | 0.97 | 1.31 | 0.12881 |
|  | **Merthyr Tydfil** | 1.09 | 0.94 | 1.27 | 0.26314 |
|  | **Monmouthshire** | 1.08 | 0.89 | 1.30 | 0.44402 |
|  | **Neath Port Talbot** | 1.62 | 1.43 | 1.84 | 0.00000 |
|  | **Newport** | 0.79 | 0.69 | 0.91 | 0.00118 |
|  | **Pembrokeshire** | 1.12 | 0.95 | 1.33 | 0.17595 |
|  | **Powys** | 1.05 | 0.88 | 1.25 | 0.59866 |
|  | **Rhondda Cynon Taff** | 1.23 | 1.09 | 1.39 | 0.00078 |
| **Swansea** | 1.49 | 1.32 | 1.68 | 0.00000 |
| **Torfaen** | 0.96 | 0.83 | 1.12 | 0.62127 |
| **Vale of Glamorgan** | 1.02 | 0.88 | 1.17 | 0.81997 |
| **Wrexham** | 1.22 | 1.06 | 1.39 | 0.00419 |
| **WIMD 2014 - overall** |  |  |  |  |
| **1 (most deprived)** | 1.67 | 1.57 | 1.77 | 0.00000 |
| **2** | 1.52 | 1.43 | 1.62 | 0.00000 |
| **3** | 1.37 | 1.29 | 1.46 | 0.00000 |
| **4** | 1.28 | 1.19 | 1.37 | 0.00000 |
| **Local area - urban/rural** |  |  |  |  |
| **Rural town** | 1.08 | 1.03 | 1.13 | 0.00277 |
| **Rural village** | 1.20 | 1.12 | 1.29 | 0.00000 |
| **No of adult in the household** |  |  |  |  |
| **1** | 1.08 | 1.04 | 1.12 | 0.00028 |
| **3** | 1.16 | 1.10 | 1.22 | 0.00000 |
| **4 or above** | 1.17 | 1.10 | 1.24 | 0.00000 |
| **No of children in the household (excluding the cohort member)** |  |  |  |  |
| **0** | 1.08 | 1.03 | 1.13 | 0.00220 |
| **2** | 1.21 | 1.16 | 1.26 | 0.00000 |
| **3** | 1.43 | 1.35 | 1.52 | 0.00000 |
| **4 or above** | 1.63 | 1.52 | 1.75 | 0.00000 |
| **Child's age in the academic year** |  |  |  |  |
|  | 0.31 | 0.30 | 0.33 | 0.00000 |
| **School session absences** |  |  |  |  |
|  | 1.02 | 1.02 | 1.02 | 0.00000 |
| **Unauthorised absences** |  |  |  |  |
|  | 1.00 | 0.99 | 1.00 | 0.00064 |

### Result from decision tree

The training model consisted of 127,090 individuals who lived in Wales (excluding testing dataset). According to the importance of the variables in the training model, the risk factors can be presented as: FSM, gender (boy), total number of absent sessions in school, child’s age while completing Foundation Phase, children with any emergency hospital admission, children with any A&E attendance, children with asthma, low birth weight, maternal substance misuse related GP record, maternal substance misuse related hospital admission, not being breastfed, children with ear problems and number of children in the household (higher number). The final decision tree model has been shown in Figure 2. Here are some case studies of the branches described in the decision tree model.

**Figure 2:**
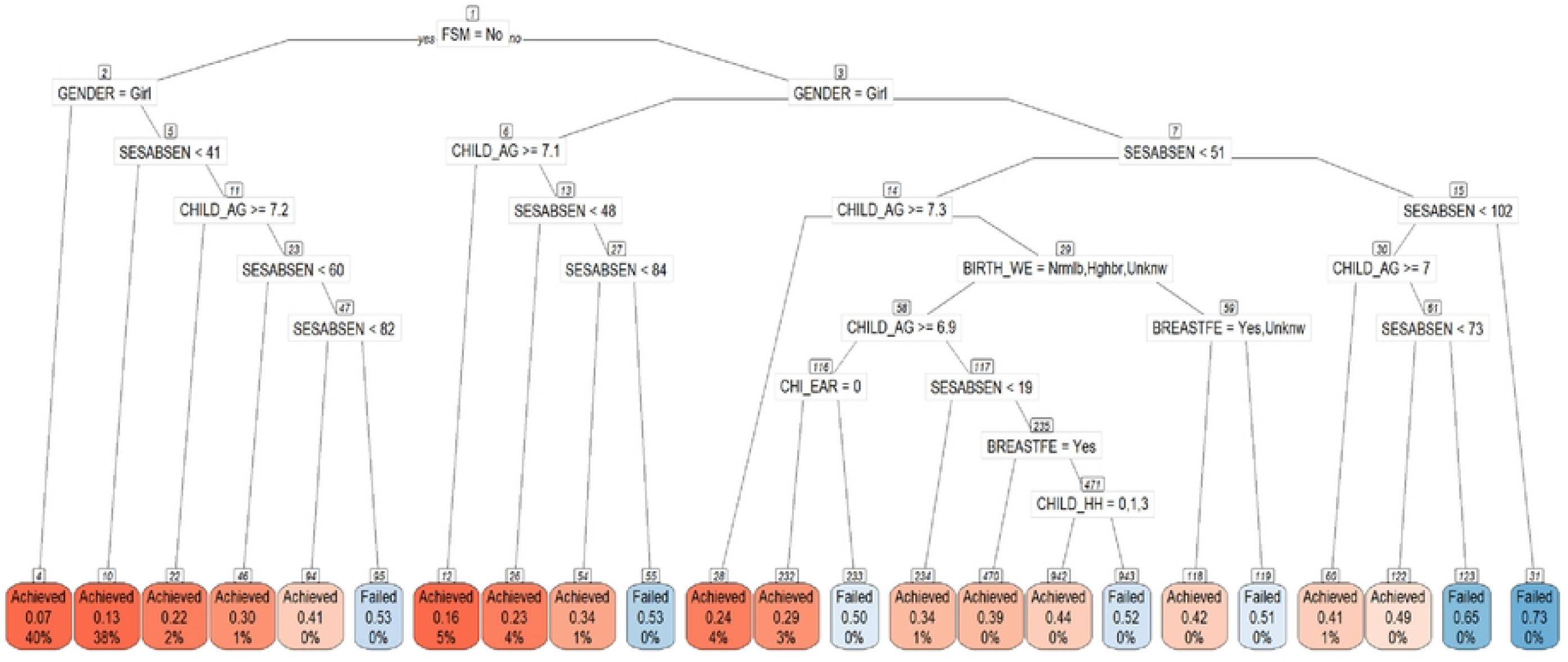
Decision tree for the children without learning difficulty. 1. IF children are eligible for FSM (higher family level deprivation) -> Gender- Boys -> Total number of absent sessions more than 102 THEN the probability of Failed is 73% (terminal node 31) and this constitutes of the 0.1% of the training model population. 2. IF children are not eligible for FSM (lower family level deprivation) -> Gender- Girls THEN the probability of Achieved is 93% (terminal node 4) and this constitutes of the 40% of the training model population. 3. IF children are not eligible for FSM (lower family level deprivation) -> Gender- Girls -> Total number of absent sessions more than 41 THEN the probability of Achieved is 87% (terminal node 10) and this constitutes of the 38% of the training model population.

There were 14,575 children in the testing dataset. The model performance has been explained with the help of a confusion matrix. The model achieves 85.21% accuracy, 4.94% sensitivity, 99.37% specificity, 58.06% positive predictive values and 85.56% negative predictive value and 15% prevalence (see Table 3 and 4).

**Table 3:** Confusion matrix/two by two table of the DT model.

| Prediction | Reference<br>(Children without learning difficulty)<br>n = 14,575 |  |
| --- | --- | --- |
|  | Did not achieve (P) | Achieved (N) |
| Did not achieve(P) | 108 (TP) | 78 (FP) |
| Achieved (N) | 2,078 (FN) | 12,311 (TN) |

**Table 4:** Prediction model performance (n= 14,575 children from Rhondda Cynon Taff)

|  | Accuracy | Sensitivity | Specificity | Positive Predictive Value | Negative Predictive Value | Prevalence |
| --- | --- | --- | --- | --- | --- | --- |
| **Decision Tree** | 85.21% | 4.94% | 99.37% | 58.06% | 85.56% | 15% |

## Discussion

This study investigated the significant risk factors associated with low school readiness and developed two holistic models on a national level routine data framework. Here the regression model helped to identify the risk factors with the highest association/Odds Ratio but might not be common or frequently observed on a population level, the decision tree on the other hand contributed to identify the most important and common/frequent risk factors. Infrequent but highly associated events/factors which affect a child’s school readiness include if the mother has a learning disability (0.14%), the child has epilepsy (0.64%) or is born extremely low birth weight (1%). However, there were also factors which were both highly associated and common such as being a boy (51%), where the odds of not being school ready is 2.11 than a girl (aOR more than twice that of girls), family level deprivation (eligible for FSM) which includes 19.5% of children, doubles the risk that they will not be school ready (aOR: 1.85). Low school attendance in early years (e.g., nursery) is associated with being 2% less likely to be school ready for every day missed in nursery.

The findings from our study suggest that rising poverty and the cost of living crisis are likely to result in poorer school readiness and poorer educational attainment. This will put a strain on school resources as children enter school. Boys being disadvantaged compared to girls has been noted in other research (25). In fact, it is suggested that family instability (separation, divorce, second families) affects boys more than girls, with a lack of a male influence impacting on behavioural difficulties (25) and that recent population increases in family instability can help explain a trend in lower attainment for boys at all levels. In addition, existing research clearly demonstrates that deprivation is a strong predictor of poor school readiness (9,26). Various indicators of deprivation such as parental employment, lower parental educational attainment, lower income, less time with the child, poorer play/local area facilities have been identified as significantly linked with low school readiness (7). Our findings such as the significant association between living in family level (eligibility for FSM) and area level (most deprived WIMD) deprivation and higher chance not to be school ready are along the similar lines reported in the literature (9,27,28). Hence, it is suggested that pre-school investment (28) and free childcare can overcome some of the risk factors associated with deprivation.

### Strengths and limitations

This study is based on linked data for an entire country over a 6-year period, and so can give a good population estimate of effects. However, it can only examine factors which are recorded using routine data. Important factors such as parenting style, time spent with the child reading, playing, and interacting cannot be captured with this data but would be important factors associated with school readiness. Another limitation of the study is that it only included the children who were in Wales during the entire study period and were removed if the children moved out of Wales as we were unable to capture their exposure records. A major strength of the study is that it incorporated birth data to build the model to identify the risk factors of low school readiness, hence these findings can be used to estimate the school readiness of a child from birth as many of these factors are present in the first years of life (gender, deprivation, gestational age, parental health) and so those at risk can be supported through access to childcare, parenting support and supporting breastfeeding. In addition, the school readiness for local children coming to a school can be predicted and this means schools can have the right resources in place to help the specific catchment of children coming to their school.

## Conclusion

This study shows that school readiness can be predicted by routine data and suggests that earlier intervention (access to childcare, mother/baby groups, community activities, parenting interventions) could help improve the outcomes for children who are at a high risk of poor school readiness. This is especially true in deprived areas with poor access to childcare and where there are child or adult health problems. It has been observed that intervention programmes like Flying Start has positive effects on the children living in deprivation including improved school attendance and better educational outcomes than their peers who are in similar condition but not under Flying Start programme (29). This work suggests that interventions which focused on boys in deprived areas, encourage or facilitated attendance in nursery in the early years, investment in early years childcare and promoting breastfeeding would have a significant impact on school readiness. Interventions such as parenting programmes which supported families with parental learning difficulties, support when there is parental or child illness (e.g., community tutoring volunteer programmes) especially for epilepsy would make a significant difference for the child’s readiness for school. This could positively influence a child’s life trajectory by strengthening foundations for lifelong learning, improving health and wellbeing outcomes throughout the life-course, and reducing education and developmental inequalities that persist.

## Data Availability

The data have been archived in the Secure Anonymised Information Linkage Databank (https://saildatabank.com/0029), it will be avilable based on request.

## Funding

This work was funded by Public Health Wales (PHW), grant number (105186). This research has been carried out as part of the ADR Wales programme of work. The ADR Wales programme of work is aligned to the priority themes as identified in the Welsh Government’s national strategy: Prosperity for All. ADR Wales brings together data science experts at Swansea University Medical School, staff from the Wales Institute of Social and Economic Research, Data and Methods (WISERD) at Cardiff University and specialist teams within the Welsh Government to develop new evidence which supports Prosperity for All by using the SAIL Databank at Swansea University, to link and analyse anonymised data. ADR Wales is part of the Economic and Social Research Council (part of UK Research and Innovation) funded ADR UK (grant ES/S007393/1).

This work was also supported by the National Centre for Population Health and Well-Being Research (NCPHWR) which is funded by Health and Care Research Wales. This work was supported by Health Data Research UK which receives its funding from HDR UK Ltd (NIWA1) funded by the UK Medical Research Council, Engineering and Physical Sciences Research Council, Economic and Social Research Council, Department of Health and Social Care (England), Chief Scientist Office of the Scottish Government Health and Social Care Directorates, Health and Social Care Research and Development Division (Welsh Government), Public Health Agency (Northern Ireland), British Heart Foundation (BHF) and the Welcome Trust.

This work uses data provided by patients and collected by the NHS as part of their care and support. Anonymised data held in the Secure Anonymised Information Linkage (SAIL) Databank has been used in this study. We would like to acknowledge all the data providers who enable SAIL to make anonymised data available for research.

## Contributorship statement

All authors contributed to the study conception and design. Initial literature review was conducted by Emily Marchant. The core dataset was prepared by Amrita Bandyopadhyay. Further data preparation and the full analysis was done by Amrita Bandyopadhyay. Hope Jones, and Michael Parker contributed to the analysis. The first draft of the manuscript was written by Amrita Bandyopadhyay and all authors commented on previous versions of the manuscript. All authors read and approved the final manuscript.

Conceptualization: Sinead Brophy, Julie Evans and Amrita Bandyopadhyay; Literature review: Emily Marchant; Methodology: Amrita Bandyopadhyay and Sinead Brophy; Formal analysis and investigation: Amrita Bandyopadhyay Writing - original draft preparation: Amrita Bandyopadhyay; Writing - review and editing: Emily Marchant, Michael Parker, Hope Jones, Julie Evans, and Sinead Brophy, Supervision: Sinead Brophy.

### Competing interest’s statement

The authors declare that they have no conflict of interest.

The views expressed in this paper are those of the authors and not necessarily those of the Office for National Statistics

### Participant consent

The study did not require participant consent as it utilises the anonymised data.

### Ethical approval

No human participants were included.

### The original protocol

Not applicable

## RECORD checklist

RECORD checklist has been added as a Supplementary file.

## Data sharing statement

The data have been archived in the Secure Anonymised Information Linkage Databank (https://saildatabank.com/0029)

## Notes

### Competing Interest Statement

The authors have declared no competing interest.

### Author Declarations

This study has included anonymised linked routine data and the study has been approved by the SAIL Databank independent Information Governance Review Panel (IGRP) (approved project number 0916 – WECC Phase 4). This is a governance review panel which has it’s ethics committee. The IGRP approval documents have been attached with this submission.

